# Characterizing features of outbreak duration for novel SARS-CoV-2 variants of concern

**DOI:** 10.1101/2022.01.14.22269288

**Authors:** Alex D. Washburne, Nathaniel Hupert, Nicole Kogan, William Hanage, Mauricio Santillana

## Abstract

Characterizing the dynamics of epidemic trajectories is critical to understanding the potential impacts of emerging outbreaks and to designing appropriate mitigation strategies. As the COVID-19 pandemic evolves, however, the emergence of SARS-CoV-2 variants of concern has complicated our ability to assess in real-time the potential effects of imminent outbreaks, such as those presently caused by the Omicron variant. Here, we report that SARS-CoV-2 outbreaks across regions exhibit strain-specific times from onset to peak, specifically for Delta and Omicron variants. Our findings may facilitate real-time identification of peak medical demand and may help fine-tune ongoing and future outbreak mitigation deployment efforts.

## Body

While emerging pandemic outbreaks may be challenging to characterize due to their novelty, their local manifestations – for example, outbreak duration – nonetheless may have common features. Identifying common features of new variant-attributable outbreaks may help decision-makers in locations where outbreaks have not been observed make the appropriate preparations to mitigate imminent outbreaks based on information obtained from previously observed outbreaks in other (comparable) locations.

Whether due to pathogen spillover from wildlife reservoirs^1^ or endogenous evolution of a novel variant,^2,3,4^ the introduction of a novel pandemic-capable infectious agent occurs in one place at one time and subsequently spreads from this locale. One approach to estimate outbreak trajectories from emergent pathogens is to collect individual-level data, such as basic reproductive numbers, serial intervals, and case severity profiles, and input these parameters into population-level compartmental models.^5^ A complementary approach that we explore leverages data from early, well-documented outbreaks as reference points for defining key features of the epidemic curve, and then continually aggregates evidence of variant-specific outbreak features or explainable variation for use in comparative epidemiological nowcasting.^6^

During the SARS-CoV-2 pandemic, global transmission has coincided with expansive viral evolution,^7^ including several punctuated equilibria producing novel and heavily mutated variants of concern with distinct epidemiological properties.^8^ The continued evolution of novel variants has produced a divergence in the slow and specialized nature of traditional model-based outbreak forecasting and the comparatively more rapid and distributed demand for assessment of the impact of novel variants on local outbreak duration, burden, and medical demand. Here, using a novel method for conducting such rapid data analysis across the US and Europe, we report a common finding across outbreak cycles driven by two novel SARS-CoV-2 variants: Delta and Omicron.

The original pandemic waves of 2020 and early 2021 were highly irregular, with accelerating and decelerating transmission impacted by combinations of immunity, human behavioral patterns, seasonal forcing, and irregularly timed interventions across regions. While there is some evidence that less-mitigated outbreaks converged to a common population fatality rate^6^, the heterogeneous seasonality, containment, mitigation, and behavioral changes around the world led to few universal patterns of outbreak cycles one could use for comparative forecasts of likely epidemic trajectories caused by the original lineage of SARS-CoV-2.

The Delta variant then caused an outbreak in India in the first half of 2021 and later caused outbreaks in the UK, South Africa, and elsewhere. In the United States, early Delta outbreaks occurred in the states of Missouri and Arkansas. Early archetypal **Delta outbreaks** in the UK, South Africa and India had similar durations, lasting roughly **75-90 days from the onset of case growth to their peaks** (FIGURE, A). Notably, the highly similar transmission trajectories of early Delta outbreaks proved useful for predicting the transmission trajectories and duration of later outbreaks across the United States (FIGURE, B).

**Figure.**
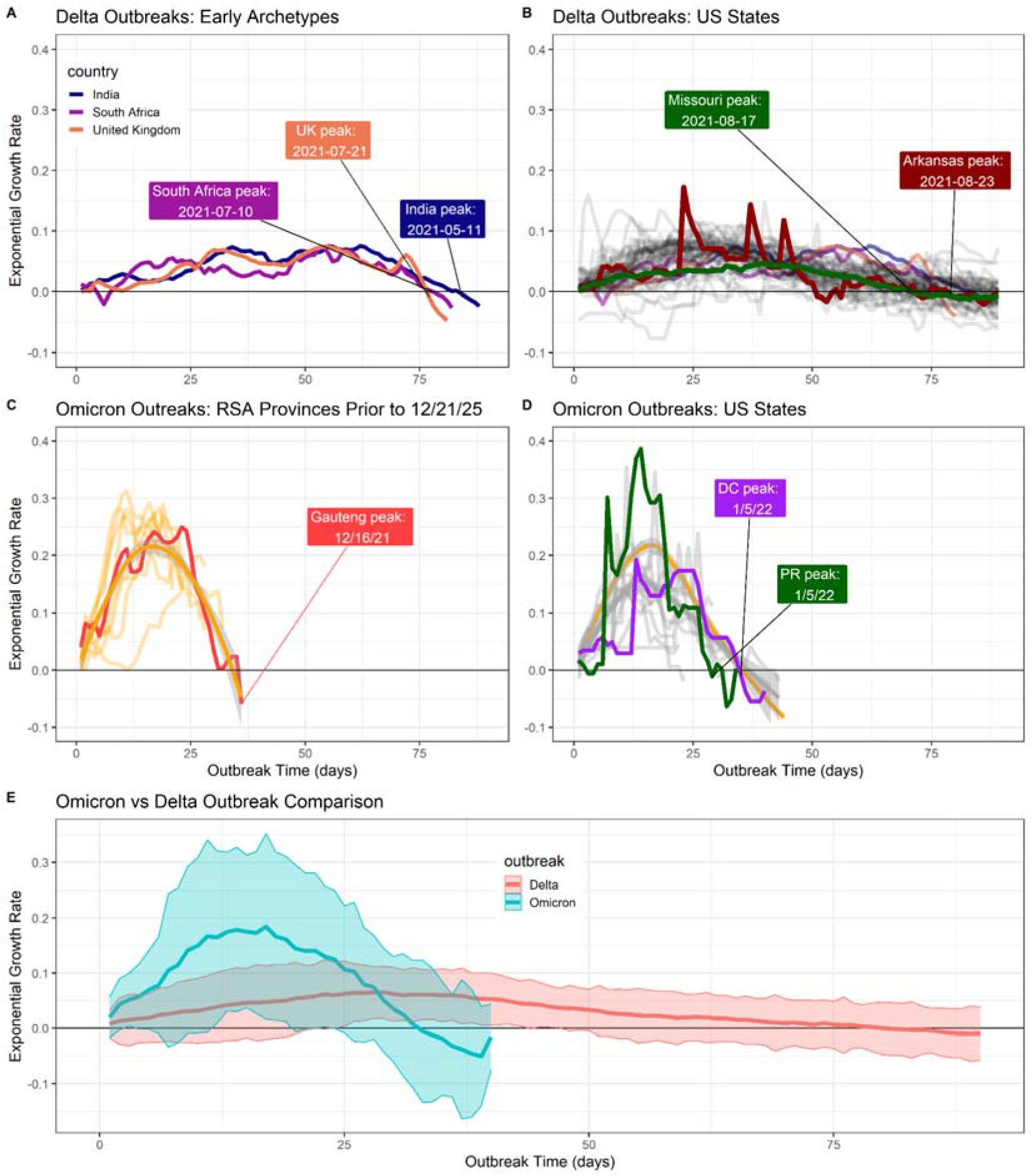
Outbreak durations for Delta and Omicron. **(A)** Early Delta outbreaks in India, UK and South Africa peaked 75-90 days after the onset of case growth. **(B)** Later US outbreaks corroborated these slow and longer-lasting outbreaks for Delta, including early US outbreaks of Missouri and Arkansas. **(C)** Omicron outbreaks in South Africa also shared a characteristic timescale, peaking 35 days after the onset of outbreaks. **(D)** Ongoing, early US Omicron outbreaks up to January 10, 2022 are corroborating the trajectory estimated from South African provinces, suggesting rapid case growth peaks approximately 35 days after outbreaks begin. **(E)** Plotting mean +/-2sd case growth rates across outbreaks, as a function of days since outbreaks began, reveals distinct trajectories of outbreaks from novel SARS-CoV-2 variants of concern.

Omicron was first reported in South Africa in November, 2021^9^ with early evidence suggesting it could cause a rapid rate of growth in cases and a higher rate of reinfections.^10^ The waning protection of vaccines against infection was documented prior to the arrival of Omicron,^11^ and further evidence suggested that Omicron’s heavily mutated spike protein lowers the binding affinity of antibodies generated by current SARS-CoV-2 vaccines and used in monoclonal antibody therapy.^12^ The complex and highly uncertain landscape of immunity against Omicron produced similar complexity and uncertainty about its outbreak trajectories outside of South Africa.

Here we present our finding that **Omicron outbreaks** across South African provinces are following a comparable pattern to earlier Delta outbreaks, albeit on a shorter timescale: South African provinces saw rapid case growth and equally rapid decelerations, with **an onset to peak time of approximately 30 days** (FIGURE, C). Early Omicron outbreaks in the United States are at present occurring in Hawaii, Florida, Puerto Rico, Washington D.C., and New York City. As of January 10, 2020 the US Omicron outbreaks appear to be following transmission trajectories akin to earlier outbreaks in South African provinces.

These early-identified universal features of Omicron outbreak trajectories and the previous efficacy of similar features in anticipating Delta outbreak trajectories produces a hypothesis that, absent highly effective containment or mitigation measures, Omicron outbreaks to date have peaked roughly one month after they start. Whether this will remain the case and be observed in all contexts is not clear, but it is a remarkably consistent observation across very different locales with very different testing and reporting practices, and distinct pandemic/vaccination histories. It should also be noted that these empirical observations do not account for population size or heterogeneity; we expect that outbreaks in smaller populations (such as small communities like colleges) and more homogeneous jurisdictions like cities may be more rapid than in larger jurisdictions like the United States as a whole or Europe, which is comprised of smaller community-level outbreaks with different start-dates. The choice of scale and heterogeneity could explain the longer duration of the Indian Delta outbreak compared to the UK, South Africa, and US states, for example. Similarly, we do not distinguish between declines due to growing population level immunity and changes in behavior, both of which likely contribute to the observed dynamics.

The duration of outbreaks caused by novel pathogens can be a useful piece of information in strategizing mitigation policies and medical logistics. Another useful piece of information will be case severity profiles, which will facilitate estimation of upper bounds on medical demand. Early evidence suggests Omicron virions are better able to replicate in the bronchus than the lungs,^13^ may have a lower risk of hospitalization per case,^14^ and may lead to shorter hospitalization times.^14^ Early population-level estimates of the determinants of Omicron burden may require controlling for the variable start-dates and transmission trajectories identified here, as, all else equal, states farther along their outbreaks can be expected to have higher medical burden.

The dynamic interplay of immunity (both vaccine- and infection-induced), strain-specific biological impact (e.g., predilection for upper vs. lower airway infection), the age structure of the unvaccinated population, and other factors will determine the magnitude and types of medical resources required for optimal patient management for Delta, Omicron, and other new COVID-19 strains. Daily data-driven comparative epidemiological analyses, similar to those described here, may allow hospital managers to approximate the duration of upcoming rises in medical demand in an ongoing, dynamic, evidence-informed fashion. Such analytic approaches should be implemented and dynamically updated to aid in local decision-making environments leveraging regional, cross-national, or global comparators. To that end, the code used to create the figure in this manuscript is available on https://github.com/reptalex/voc_outbreak_time.

## Methods

### Data source and growth rate estimation

Data were downloaded daily using the R package COVID-19.^15^ The exponential growth rates of cases in region *i* at time *t, r(i,t)*, were estimated by first removing outliers and then computing the filtering density of a negative binomial state-space model with day-of-week fixed effects, described in greater detail elsewhere.^6^ The filtering density was used to ensure all historical comparisons of outbreaks at different points of progress are incorporating information available up until that point, avoiding future-peeking effects of smoothing. Holidays and their subsequent delayed reporting were removed for US states.

The day-of-week case reporting patterns changed in many US states between the Delta and Omicron waves. To account for this, the US state growth rates for Delta were estimated starting on 2021-04-01; for Omicron, growth rate estimation started 21 days prior to the onset of each state’s Omicron wave.

### Outbreak start-dates

Across all regions described here, Delta outbreaks began after a period of declining cases, and consequently the start date of Delta outbreaks were defined as the dates of the local minimum of expected new cases under the filtering density falling within a specified date range. Date ranges were chosen to crop out well-known Alpha outbreaks. The start date for Delta was estimated to be 2021/02/17 in India, 2021/04/24 in South Africa, and 2021/05/07 in the UK.

Omicron was reported to the world by South African authorities on November 23, 2021. Omicron outbreaks in South Africa similarly followed a period of declining caseloads, and so the start-date of Omicron outbreaks in South Africa was defined as the first date after November 11^th^, which was then followed by at least 10 days of sustained growth in cases.

In US states, Omicron outbreaks occurred during probable seasonal forcing of COVID-19 transmission when cases were growing in many states. Consequently, we could not use the onset of case growth to define start-dates across US states nor is there widespread genomic surveillance permitting start-dates defined by strain composition. To estimate effective start-dates of Omicron outbreaks during ongoing transmission of another variant, we defined start-dates of Omicron in the US using the characteristically rapid growth of Omicron. Any state with case growth rates exceeding r>0.125 were declared as Omicron outbreaks, and their start dates were the latest preceding date with r>0.02.

## Data Availability

All data and scripts are available online at https://github.com/reptalex/voc_outbreak_time

## Citations

1 Plowright RK, Parrish CR, McCallum H, et al. Pathways to zoonotic spillover. Nat Rev Microbiol. 2017;15(8):502–510. doi:10.1038/nrmicro.2017.45

2 He W, Zhang W, Yan H, et al. Distribution and evolution of H1N1 influenza A viruses with adamantanes-resistant mutations worldwide from 1918 to 2019. J Med Virol. 2021;93(6):3473–3483. doi:10.1002/jmv.26670

3 Mlcochova P, Kemp SA, Dhar MS, et al. SARS-CoV-2 B.1.617.2 Delta variant replication and immune evasion. Nature. 2021;599(7883):114–119. doi:10.1038/s41586-021-03944-y

4 He X, Hong W, Pan X, Lu G, Wei X. SARS-CoV-2 Omicron variant: Characteristics and prevention [published online ahead of print, 2021 Dec 16]. MedComm (2020). 2021;2(4):838–845. doi:10.1002/mco2.110

5 Tolles J, Luong T. Modeling Epidemics With Compartmental Models. JAMA. 2020;323(24):2515–2516. doi:10.1001/jama.2020.8420

6 Washburne A, Silverman J, Lourenco J, Hupert N. Analysis and visualization of epidemics on the timescale of burden: derivation and application of Epidemic Resistance Lines (ERLs) to COVID-19 outbreaks in the US. medRxiv. 2021;05.03.21256542. doi: 10.1101/2021.05.03.21256542

7 Hadfield J, Megill C, Bell SM, et al. Nextstrain: real-time tracking of pathogen evolution. Bioinformatics. 2018;34(23):4121–4123. doi:10.1093/bioinformatics/bty407

8 Tao K, Tzou PL, Nouhin J, et al. The biological and clinical significance of emerging SARS-CoV-2 variants. Nat Rev Genet. 2021;22(12):757–773. doi:10.1038/s41576-021-00408-x

9 Viana R, Moyo S, Amoako D, et al. Rapid epidemic expansion of the SARS-CoV-2 Omicron variant in southern Africa [published online ahead of print, 2022 Jan 7]. Nat Research Briefings. 2022. doi: 10.1038/d41586-021-03832-5

10 Pulliam JRC, van Schzlkwyk C, Govender N, et al. Increased risk of SARS-CoV-2 reinfection associated with emergence of the Omicron variant in South Africa. medRxiv. 2021; 11.11.21266068. doi: 10.1101/2021.11.11.21266068

11 Goldberg Y, Mandel M, Bar-On YM, et al. Waning Immunity after the BNT162b2 Vaccine in Israel. N Engl J Med. 2021;385(24):e85. doi:10.1056/NEJMoa2114228

12 Wu L, Zhou L, Mo M, et al. SARS-CoV-2 Omicron RBD shows weaker binding affinity than the currently dominant Delta variant to human ACE2. Signal Transduct Target Ther. 2022;7(1):8. Published 2022 Jan 5. doi:10.1038/s41392-021-00863-2

13 Diamond M, Halfmann P, Maemura T, et al. The SARS-CoV-2 B.1.1.529 Omicron virus causes attenuated infection and disease in mice and hamsters. Preprint. Res Sq. 2021;rs.3.rs-1211792. Published 2021 Dec 29. doi:10.21203/rs.3.rs-1211792/v1

14 Wolter N, Jassat W, Walaza S, et al. Early assessment of the clinical severity of the SARS-CoV-2 Omicron variant in South Africa. medRxiv. 2021; 12.21.21268116. doi: 10.1101/2021.12.21.21268116

15 Guidotti E, Ardia D. COVID-19 data hub. Journal of Open Source Software. 2020 Jul 10;5(51):2376.

